# Pervasive Intestinal Carriage with Multiple Species of Extended Spectrum Cephalosporin-Resistant Enterobacterales in Children Admitted for Severe Acute Malnutrition at a Tertiary Hospital in Malawi

**DOI:** 10.1101/2025.05.30.25328637

**Authors:** Thomas Holowka, Arrington Ashford, Trieu-Vi Khuu, Alisher Bimagambetov, Courtney N. Dial, Alexander Kondwani, Amazing-Grace Tepeka, Kevin Alby, Jonathan J. Juliano, Anthony J. Garcia-Prats, Tisungane Mvalo, Bryan J. Vonasek, Emily J. Ciccone, Luther A. Bartelt

## Abstract

Using culture-based screening of stool specimens, 80.3% of 188 children hospitalized for severe acute malnutrition in Malawi carried extended spectrum cephalosporin resistant Enterobacterales (ESCR-E) at admission. 38.3% of children were colonized with multiple ESCR-E species. Over half of the colonized children did not have recent antibiotic exposure or prior hospitalization.

## Introduction

Antimicrobial resistant bacteria, such as the Extended Spectrum Cephalosporin-Resistant Enterobacterales (ESCR-E) *Klebsiella pneumoniae* and *Escherichia coli*, are a growing global health threat. Despite known increases in the prevalence of ESCR-E infections and ESCR-E infection-related mortality in Sub-Saharan Africa (SSA), there is little data to inform empiric antibiotic treatment recommendations at the local level [1, 2]. This evidence gap is of particular importance in regions where pediatric severe acute malnutrition (SAM) is prevalent as SAM is known to increase risk for bacteremia and infection-related mortality [3, 4]. Recent reports showing that up to 40% of *E. coli* and 85% of *K. pneumoniae* clinical isolates from children in SSA are ESCR highlight the pressing need to assess the current prevalence of ESCR-E infection in children with SAM in the community [5].

We recently reported on a cohort of children admitted for SAM at Kamuzu Central Hospital (KCH) in Lilongwe, Malawi, in which 43% of all Gram-negative organisms and 35% of *E. coli* isolated from admission blood and urine cultures were ESCR [6]. Since ESCR-E establish colonization in the intestine prior to causing invasive infection, we screened fecal samples from these children to determine prevalence of ESCR-E carriage upon admission to the hospital.

## Methods

We performed a cross-sectional screen for intestinal ESCR-E colonization in stool samples available from children admitted with SAM. The primary study was a prospective observational cohort of 212 children 6-59 months old with complicated SAM admitted to KCH from February 2023 to January 2024. The study protocol, eligibility criteria, consent processes, study activities, and data collection have been described previously [6]. These included collection of routine blood and urine cultures at admission and records of recent antibiotic use (within the previous 5 days) and any prior hospitalizations as reported by the participants parent/guardian. The first available stool was collected for stool culture within 24 hours of admission, and residual samples following initial stool pathogen testing on site were stored at -80?. Specimens were then shipped on dry ice to the University of North Carolina, Chapel Hill, NC, USA for further testing.

ESCR-E screening was performed using culture enrichment in antibiotic selective media. Scrapings of frozen fecal specimens were inoculated into tryptic soy broth supplemented with cefotaxime (1 µg/mL) and incubated at 37? for 20 hours. Broth cultures were streaked onto MacConkey agar supplemented with cefotaxime (1 µg/mL) then incubated at 37? for 20 hours. Resulting isolates were examined for unique colony morphologies, defined according to colony size, shape, color, and texture. Species identification was performed on all unique isolates using Matrix Assisted Laser Desorption Ionization Time of Flight (MALDI-TOF) with the bioMerieux VITEK Mass Spectrometer according to manufacturer’s instructions. Isolates identified as a member of Enterobacterales were considered ESCR-E. Antimicrobial susceptibility testing (AST) was performed on all isolates using Kirby-Bauer disk diffusion on Mueller-Hinton Agar following standard protocols [7]. Resistance cut-offs were determined according to 2025 Clinical and Laboratory Standards Institute cut-offs listed in the CLSI M100 35^th^ edition [8]. Organism/drug combinations that tested as intermediate were included as non-susceptible for purposes of analysis.

We used standard summary statistics to describe stool testing and AST results. Unadjusted prevalence ratios (PR) with confidence intervals (CI) were determined for comparison of colonization rates in patients with or without prior hospitalization or antibiotic exposure using Stata 19 software. Graphical representations were created with GraphPad Prism 10 Software.

## Results

Stool specimens were available from 188 unique participants, and all specimens underwent intestinal colonization screening. ESCR-E were recovered from 80.3% (151/188) of participants. A total of 293 morphologically distinct colonies were isolated and 46.8% were identified as *E. coli*, 36.9% as *K. pneumoniae* and 11.9% as *Enterobacter* species (Figure 1A). Multiple morphotypes in stool specimens were detected in 53.2% (100/188) of total participants, including 66.2% (100/151) of ESCR-E screen-positive participants. Multiple species of ESCR-E were identified in stool specimens from 38.3% (72/188) of participants, including 47.7% (72/151) of ESCR-E screen-positive participants (Figure 1B). Across all stool specimens, 54.3% grew *E. coli*, 46.8% grew *K. pneumoniae*, 15.4% grew *Enterobacter spp*., and 2.7% grew other Enterobacterales. Non-Enterobacterales organisms (*Acinetobacter baumannii* and *Pseudomonas aeruginosa*) were isolated from 3.8% of the specimens (Figure 1C). Kirby-Bauer disc diffusion confirmed ceftriaxone non-susceptibility in 100% of cefotaxime-broth enrichment recovered isolates. Non-susceptibility to multiple antibiotic classes was observed in most of the isolates including gentamicin (79.2%), ciprofloxacin (62.6%) and chloramphenicol (26.6%). Non-susceptibility to other beta-lactams was also detected including ampicillin (100%), cefepime (40.5%), piperacillin-tazobactam (21.1%) and ertapenem (6.2%) (Figure 1D, Table S1).

**Figure 1:**
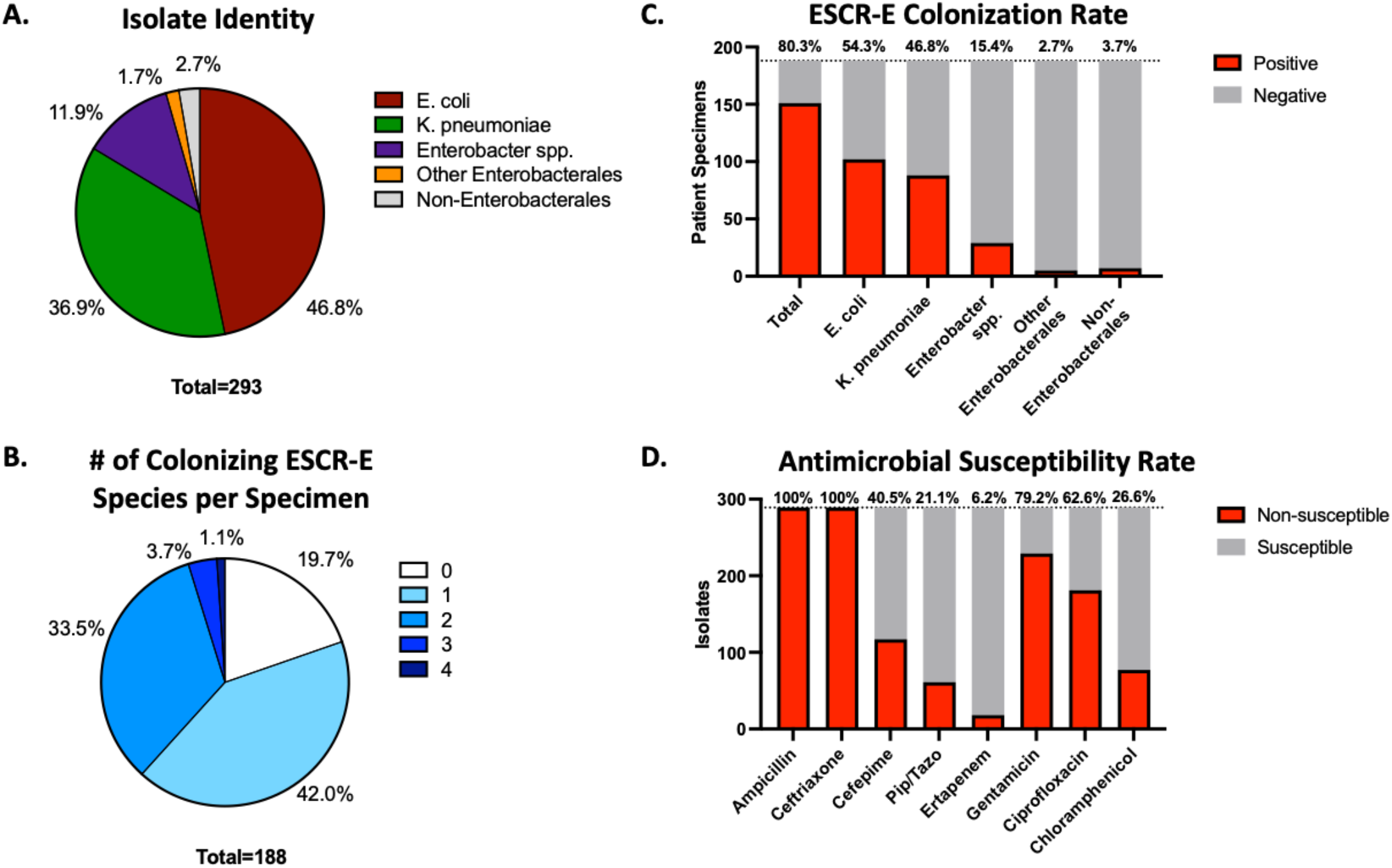
ESCR-E colonization rate, species identification and antimicrobial susceptibility. (A) Identification of morphologically distinct isolates performed via MALDI-TOF. Other Enterobacterales include *Klebsiella oxytoca* (n=3) *Erwinia bilinigial* (n=1), *Citrobacter freundii* (n=1). Non-Enterobacterales include *Acinetobacter baumanii* (n=5) *Pseudomonas aeruginosa* (n=2). (B) Number of distinct species isolated from each individual stool specimen. (C) Number of individual specimens with growth on ESCR-E screen with total number and percentage positive for individual species. (D) Number of isolates and percentage of total tested (n=207) susceptible to each individual antimicrobial tested.

A minority of patients were reported to have recent antibiotic exposure within the previous 5 days (72/188, 38.3%) or any prior hospitalization (78/188, 41.5%). ESCR-E colonization was similar in participants regardless of antibiotic exposure within the prior 5 days, with 84.7% (61/72) colonized in antibiotic-exposed versus 77.6% (90/116) colonized in antibiotic-unexposed (PR = 1.09, 95% CI: 0.95-1.25). Colonization was also similar for participants with any prior hospitalization (85.9%; 67/78) versus those without prior hospitalization (76.4%; 84/110) (PR = 1.12; 95% CI: 0.98-1.29). Notably, all 11 participants in the cohort with a possible ESCR-E organism (Gram negative organism non-susceptible to 3^rd^-generation cephalosporins) grown from an admission clinical culture (3 with positive blood cultures, 7 with positive urine cultures, 1 with positive blood and urine cultures) had concomitant ESCR-E colonization. In total 7.3% (11/151) of ESCR-E colonized participants had admission clinical culture positivity for possible ESCR-E versus 0% (0/37) of participants without colonization.

## Discussion

Africa has the highest antimicrobial resistance-related mortality rate (348.3 per 100,000) of any WHO global region, and over a quarter of these deaths are due to ESCR-E [2]. ESCR-E infection rates have been steadily rising in children in SSA, encompassing a majority of bloodstream non-*Salmonella* Enterobacterales isolates in a retrospective study of children admitted to a hospital in Blantyre, Malawi[9]. Our findings in a cohort of children with SAM at a tertiary care hospital in Lilongwe, Malawi, demonstrate pervasive colonization with ESCR-E (80.3%) in children at the time of admission. This is a much higher ESCR-E colonization rate than the 42% colonization rate recently reported among community-dwelling adults in Blantyre, Malawi [10]. To our knowledge, this is the highest rate of colonization at admission reported in any pediatric cohort in SSA. For reference, a recent review and systematic meta-analysis including 35 studies between 2009 and 2022 found an overall prevalence of ESCR-E colonization of 32.2% in children from countries across SSA, with the highest colonization rates around 60% in individual studies [11].

In addition to the extremely high colonization rate that we observed, most children in this cohort were colonized with multiple morphologically distinct isolates of ESCR-E, including simultaneous colonization with different ESCR-E species and potentially multiple strains of the same species. There is limited literature regarding ESCR-E co-colonization in children in SSA, and we found reported rates of ESCR-E co-colonization ranging from 0% in a cohort of 161 children living with HIV in Ethiopia (20% ESCR-E mono-colonization) to as high as 12.4% co-colonization in cohort of 603 children >2 years of age in Tanzania (34.3% overall ESCR-E colonization) [12, 13]. In comparison, we found 38.3% colonization with multiple ESCR-E species, accounting for nearly half of all ESCR-E colonized children in our study. The presence of multiple ESCR-E species together with a very high rate of colonization with multiple ESCR-E morphotypes of the same species suggests a high density of ESCR-E organisms circulating in this particularly vulnerable population.

Our findings also suggest high community prevalence and circulation of ESCR-E in children with SAM rather than primarily through hospital acquisition. Specifically, we noted high rates of ESCR-E colonization in this cohort regardless of prior hospitalization or recent antimicrobial use, which reinforces the need for better understanding of community spread of ESCR-E in Lilongwe, and how previously unexplored risk factors like malnutrition may be independently implicated.

This analysis focused on children with SAM, and there is evidence that malnutrition can facilitate colonization with intestinal pathogens [4]. However, this study was not designed to compare ESCR-E colonization to a control cohort with normal nutritional status or less severe malnutrition, and additional studies are needed to determine if SAM or other types of malnutrition are independent risk-factors for ESCR-E colonization in children and adults.

Regardless, these findings have urgent implications for the empiric treatment of malnourished children at KCH and potentially elsewhere in SSA. ESCR-E intestinal colonization is a risk factor for invasive ESCR-E infection. Current WHO guidelines recommend that treatment of SAM include empiric antimicrobials, particularly with beta-lactam agents such as amoxicillin and ceftriaxone [14], agents that would be insufficient to cover common bacteria colonizing these malnourished children. Furthermore, we noted high rates of resistance to other antimicrobials including gentamicin which is typically used as local first-line treatment.

In summary, this study highlights the critical need for ongoing ESCR-E and antimicrobial resistance surveillance, especially in parts of SSA where persistently high rates of childhood malnutrition may serve to further increase already unacceptably high rates of resistance-related mortality.

## Data Availability

All data produced in the present study are available upon reasonable request to the authors

## Author Contributions

Specimen Processing and Data Generation: TH, AA, TVK, AB, CD, KA Specimen and Clinical Data Acquisition: AK, AGT, JJJ, AJG, TM, BJV Data Analysis: TH, EJC, LAB Manuscript preparation and critical revision: TH, KA, AJG, TM, BJV, EJC, LAB

---

**Table S1:** Antimicrobial non-susceptibility by species. Percent non-susceptibility reported.

|  | Ampicillin | Ceftriaxone | Cefepime | Piperacillin-Tazobactam | Ertapenem | Gentamicin | Ciprofloxacin | Chloramphenicol |
| --- | --- | --- | --- | --- | --- | --- | --- | --- |
| E. coli (n=137) | 100 | 100 | 52.6 | 23.4 | 6.6 | 71.5 | 70.8 | 20.4 |
| K. pneumoniae (n=108) | 100 | 100 | 33.6 | 21.5 | 0.9 | 87.9 | 49.5 | 19.6 |
| Enterobacter species (n=35) | 100 | 100 | 15 | 10 | 5 | 75 | 70 | 10 |
| K. oxytoca (n=3) | 100 | 100 | 0 | 0 | 0 | 66.7 | 66.7 | 66.7 |
| C. freundii (n=1) | 100 | 100 | 0 | 0 | 0 | 100 | 100 | 100 |
| E. bilingial (n=1) | 100 | 100 | 100 | 100 | 0 | 100 | 100 | 0 |
| A. baumannii (n=5) | 100 | 100 | 20 | 40 | 100 | 20 | 20 | 100 |
| P. aeruginosa (n=2) | 100 | 100 | 0 | 0 | 50 | 50 | 0 | 100 |
| Total (n=293) | 100 | 100 | 40.5 | 21.1 | 6.2 | 79.2 | 62.6 | 26.6 |

